# LeMeDISCO: A computational method for large-scale prediction & molecular interpretation of disease comorbidity

**DOI:** 10.1101/2021.06.28.21259559

**Authors:** Courtney Astore, Hongyi Zhou, Jeffrey Skolnick

**Author notes:** These authors contributed equally to this work (Courtney Astore, Hongyi Zhou).

## Abstract

Often different diseases tend to co-occur (i.e., they are comorbid), which yields the question: what is the molecular basis of their coincidence? Perhaps, common proteins are comorbid disease drivers. To understand the origin of disease comorbidity and to identify the essential proteins and pathways underlying comorbid diseases, we developed **LeMeDISCO** (**L**arge-Scal**e M**olecular Int**e**rpretation of **Dis**ease **Co**morbidity), an algorithm that predicts disease comorbidities from shared mode of action (MOA) proteins predicted by the AI-based **MEDICASCY** algorithm. **LeMeDISCO** was applied to predict the general occurrence of comorbid diseases for 3608 distinct diseases. To illustrate **LeMeDISCO**’s power, we elucidate the possible etiology of coronary artery disease and ovarian cancer by determining the comorbidity enriched MOA proteins and pathways and suggest hypotheses for subsequent scientific investigation. The **LeMeDISCO** web server is available for academic users at: http://sites.gatech.edu/cssb/LeMeDISCO.

## Introduction

Disease comorbidity, the cooccurrence of distinct diseases is an interesting medical phenomenon. For example, individuals having one autoimmune condition likely develop another. Interestingly, rheumatoid arthritis, autoimmune thyroiditis, and insulin-dependent diabetes mellitus cooccur, but rheumatoid arthritis and multiple sclerosis do not^1^. Previously, there have been several efforts to investigate the molecular features responsible for human disease comorbidities^2-8^. Some studies focused on particular subsets of diseases^3^ or ethnic groups, while others investigated the entire human disease network^4-7^. For example, Zhou et. al.^5^ applied text mining to search the literature for disease-symptom associations. They then predicted the entire human disease-disease network based on a calculated symptom similarity score. While this approach covers almost all human diseases, it only explains one phenotype (disease) by another phenotype (symptom). Menche *et. al*. ^6^ utilized known disease-gene associations from GWAS^9^ and OMIM combined with a protein-protein interaction network to identify connected disease gene clusters or modules. Another study also utilized known disease-gene associations and protein-protein interaction networks to characterize disease-disease relationships without requiring gene clusters^7^; thus, its disease coverage is better than in ref.^6^. A deeper analysis of these studies, which have low recall rates, demonstrates that focusing entirely on shared genes is insufficient to predict disease comorbidity or identify its origins. They miss collective effects arising from both direct and indirect protein-protein interactions and pathway correlations.

Existing studies that use known disease-gene associations are limited by data availability. Indeed, only a small fraction of diseases has known associated genes. For example, ref.^7^ only covers 1,022 of the 8,043 diseases in the Disease Ontology database^10^, with just 6,594 pairs of diseases having a non-zero number of shared genes. Similarly, ref.^6^ found that most (59%) of their 44,551 disease pairs do not share any genes. To address these limitations, we developed **LeMeDISCO** which extends our recently developed **MEDICASCY** machine learning approach^11^ for predicting disease indications and mode of action (MOA) proteins (as well as small molecule drug side effects and efficacy) to predict disease comorbidities and the proteins and pathways responsible for their comorbidity. We then show that **LeMeDISCO** covers a broader spectrum of comorbid diseases than existing approaches. Assuming that the most enriched comorbid proteins are responsible for disease comorbidity, we determine the most frequent comorbidity enriched MOA proteins. These proteins are then employed in pathway analysis^12^. As examples, we predict the comorbid diseases, comorbidity enriched MOA proteins, and pathways associated with coronary artery disease (CAD) and ovarian cancer (OC).

## Results

### Benchmarking Results of LeMeDISCO

To assess its relative performance, we compared the results of **LeMeDISCO** to three other methods, the XD score^7^, the S_AB_ score and the Symptom Similarity score^5^. Table 1 summarizes the results. We define a positive comorbidity pair when their log(RR) > 0, XD score > 0, *S*_*AB*_ score < 0, or the symptom similarity score > 0.1^13^ and p-value < 0.05 for J-score. The relative risk RR is defined in eq. 4a and is the probability that two diseases occur in a single individual relative to random. The φ**-**score is the Pearson’s correlation for binary variables and is defined in eq. 4b. Mapping the DOIDs from the Human Disease Ontology database to the ICD-9 IDs of ref.^14^, we obtain 198,149 disease pairs for use in **LeMeDISCO** benchmarking. All correlations of the J-score with the log(RR) score and φ**-**score are statistically significant (p-value < 0.05).

**Table 1.** Comparison of **LeMeDISCO**’s J-score with the XD score, NG, *S*_*AB*_ score and symptom similarity for correlations with comorbidity quantified by the log(RR) score, φ**-**score and recall^a^.

|  | Unbinned<sup>b</sup> |  | 10 bins<sup>b</sup> |  | Recall |
| --- | --- | --- | --- | --- | --- |
| | Log(RR) score | ϕ-score | Log(RR) score | ϕ-score | |
| 198,149 disease pairs<sup>c</sup> |  |  |  |  |  |
| LeMeDISCO | **0.312(0.0)** | **0.218(0.0)** | **0.933(8.1x10<sup>-5</sup>)** | **0.900(3.9x10<sup>-4</sup>)** | **49.7%** |
| 29,783 pairs<sup>d</sup> |  |  |  |  |  |
| LeMeDISCO | **0.185(0.0)** | **0.138(0.0)** | **0.939(5.6x10<sup>-5</sup>)** | **0.829(3.0x10<sup>-3</sup>)** | **56.0%** |
| XD score<sup>7</sup> | 0.050(5.9x10<sup>-18</sup>) | 0.082(0.0) | 0.445(0.20) | 0.252(0.48) | 6.5% |
| NG<sup>e</sup> | 0.008(0.17) | 0.058(1.3x10<sup>-23</sup>) | -0.436 | -0.175 | - |
| 947 disease pairs<sup>f</sup> |  |  |  |  |  |
| LeMeDISCO | **0.217(1.5x10<sup>-11</sup>)** | **0.282(9.0x10<sup>-19</sup>)** | **0.682(0.030)** | **0.688(0.028)** | **75.8%** |
| $S_{AB}$ score<sup>6</sup> | -0.188(5.5x10<sup>-9</sup>) | -0.218(1.2x10<sup>-11</sup>) | -0.671(0.034) | -0.473(0.17) | 8.5% |
| 2,630 disease pairs<sup>g</sup> |  |  |  |  |  |
| LeMeDISCO | 0.184(1.9x10<sup>-21</sup>) | **0.196(3.5x10<sup>-24</sup>)** | 0.774(8.6x10<sup>-3</sup>) | 0.654(0.040) | 71.1% |
| Symptom similarity<sup>5</sup> | **0.337(0.0)** | **0.197(1.6x10<sup>-24</sup>)** | **0.950(2.6x10<sup>-5</sup>)** | **0.960(1.1x10<sup>-5</sup>)** | **100%** |

<sup>a</sup> Numbers in parentheses are the p-values of the corresponding correlation. Bold indicates the best results for the given data set.
<sup>b</sup> Unbinned means raw data; each pair is a data point. 10 bins: partitioning the prediction scores into 10 equal size bins. In each bin, the log(RR) & $\phi$ -score are averaged over data points in the bin. This gives equal weight to the rare prediction scores in the correlation analysis.
<sup>c</sup> Mapping the DOID IDs from the human DO database to ICD9 IDs of Ref.<sup>14</sup>, gives a set of 198,149 disease pairs
<sup>d</sup> Mapped the ICD9 disease code to our DOID of DO and obtained a consensus subset of 29,783 disease pairs from Table 1's dataset of 97,665 disease pairs in Ref.<sup>7</sup>.
<sup>e</sup> NG is the number of shared genes between disease pairs in Ref.<sup>7</sup>.
<sup>f</sup> Consensus set of 947 disease pairs from the dataset of Ref.<sup>6</sup> and our dataset of 198,149.
<sup>g</sup> A consensus dataset of 2,630 disease pairs was obtained from their Supplementary dataset 4 of Ref.<sup>5</sup> compared to our set of 198,149 pairs.

To compare **LeMeDISCO** to the XD score, we mapped their ICD-9 disease code to the DOIDs and obtained a subset of 29,783 pairs from their dataset of 97,665 pairs^7^. As shown in Figure S1, their NG score (the number of shared genes) essentially has no significant correlation with log(RR) and only shows a correlation with the φ**-**score for unbinned data. When the data are binned, both the XD score and NG score lack significant correlations. J-score has much better correlations and recall rate than XD score.

For comparison with the *S*_*AB*_ score^6^, the MeSH^15^ disease names^16^ were mapped to their DOIDs. A consensus set of 947 disease pairs from their dataset and ours was obtained. As shown in Figure S2 and Table 1, compared to *S*_*AB*_ ^6^, for the 947 disease pairs, **LeMeDISCO**’s J-score is better than the *S*_*AB*_ score for both unbinned and binned data, and it has a much better recall rate than the *S*_*AB*_ score. The *S*_*AB*_ score has no significant correlation for the binned φ**-**score data. The reason for the worse performance of *S*_*AB*_ for binned data is as follows: unbinned data for *S*_*AB*_ are dominated by cases where no disease comorbidity is predicted (*S*_*AB*_>0), whereas for binned data, both *S*_*AB*_<0 and *S*_*AB*_>0 is important.

Next, a common dataset of 2,630 disease pairs was obtained for comparison with the symptom similarity score. As shown in Figure S3, the symptom similarity score has a better correlation than **LeMeDISCO** for the log(RR) and is almost identical to the φ**-**score for unbinned data. However, the symptom similarity score only explains the relationship of one phenotype (symptom) to another phenotype (disease). Nevertheless, all correlations of the J-score are statistically significant, and its recall rate is close to 70%.

The advantage of the J-score over the symptom similarity score is that it has a clear molecular interpretation. Moreover, **LeMeDISCO** does not rely on prior knowledge or symptomatic information. Hence, it provides a much larger coverage of comorbidity predictions for the 198,149 disease pairs, each ranked by its J-score and the corresponding p-value to reflect the statistical significance. The correlation of the J-score and the log(RR) and φ**-**score for this large set of disease pairs is shown in Figure S4.

### MEDICASCY based MOA protein prediction

The ICD-10 main classification coverage of the 3,608 diseases is shown in Figure 1A. We first examine the number of predicted MOA proteins per indication from **MEDICASCY**^11^. Using a p-value cutoff of 0.05 and including protein isoforms, the average (median) number of MOA proteins per indication is 1,744.5 (656); the maximal and minimal values are 17,324 (almost half of the total 32,584 screened proteins) for mast cell sarcoma and 0 for esophageal atresia. The histogram of the number of MOAs is shown in Figure 1B. 68.0% (41.1%) of indications have > 500 (1,000) MOA proteins; several are likely false positives or have a minor contribution to the disease, at best. However, these associations allow us to expand the protein repertoire that might be associated with each disease and are like the statistics from GWAS studies. Below, we describe the use of **LeMeDISCO** to predict disease comorbidities as well as prioritize these proteins.

**Figure 1.**
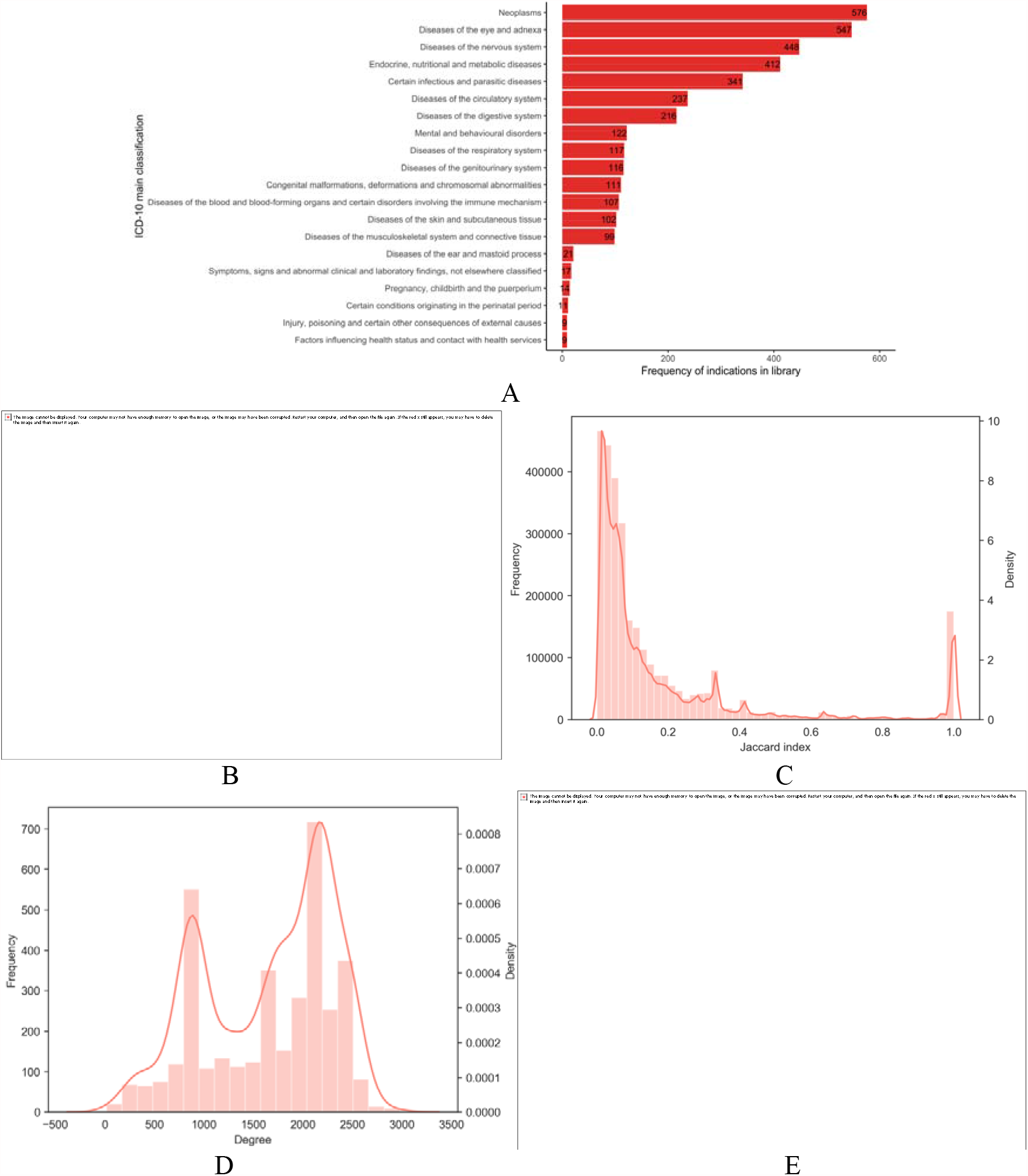
**A**. ICD-10 main classification coverage across the 3,608 diseases. Some diseases are found in multiple groups; they were counted in each group for which they are associated. **B**. Histogram of the number of MOAs. **C**. Frequency and density of the J-score for the _∼_3 million significant (p-value < 0.05), non-redundant disease pairs. **D**. Frequency and density of the degree (number of edges) of each disease (node). **E**. Fraction of diseases in the giant component of disease-disease network versus the J-score cutoff.

### Shared MOA proteins explain disease comorbidity by way of disease-disease relationships

We next examine the overall characteristics of the predicted comorbidity network of 3,608 diseases. There are a total of 6,508,832 possible pairwise disease associations. From this, there are 5,987,682 significant pairwise disease associations excluding the diagonals and 3,009,095 significant non-redundant pairwise disease associations given by **LeMeDISCO**. Only one disease, esophageal atresia, did not have any significant comorbidities predicted. Thus, 3,607 diseases contained significant comorbidities (p-value < 0.05). The density and frequency of the J-score for the significant pairs is in Figure 1C, and the density and frequency of the degree (number of edges) for each node (disease) is represented in Figure 1D. Using a p-value cutoff of 0.05, the average (median) number of comorbidities per disease is 1,650.7 (1806). The largest (smallest) number of comorbidities is 2,958 for gastric mucosal hypertrophy (17 for Canavan disease). Thus, the disease network is very dense.

The cumulative distribution for the J-score and p-values for all of the comorbidities and the top 100 are shown in Figure S5 and S6, respectively. The summary statistics of the scores for these thresholds are shown in Table S1. What is clear from these figures and Table S1, particularly for the top 100 ranked comorbidities, is that the 98.5% top ranked 100 comorbidities have a p-value < 0.005. In other words, while a p-value threshold of 0.05 is used, in reality the actual p-values employed for subsequent analysis are far more significant.

Around 46% of the disease pairs have a p-value < 0.05. This result is consistent with the ∼50% recall of large scale benchmarking (see Table 1) and the observed comorbidity from Medicare insurance claim data that 78.8% of the total 3,634,744 disease pairs have an RR > 1^14^. As shown in Figure 1E, the giant component (GP) of the disease-disease network covers the entire network when the J-score is < 0.1 and the p-value < 0.05, i.e., starting from any disease, one can walk to any other disease on the network. As the J-score cutoff increases, the number of diseases in the giant component decreases; however, the decrease is very slow. The rapid decrease only happens around a 0.45 J-score corresponding to an average p-value ∼3.6×10^−30^. Thus, the disease network is not only dense, but it is also strongly and highly significantly connected.

### LeMeDISCO identified MOA proteins

In addition to the comorbidity predictions, **LeMeDISCO** also identifies comorbidity enriched MOA proteins. The comorbidity enriched MOA proteins are hierarchically ranked by their CoMOAenrich score (defined in the Methods section). Comparing the top 100 comorbidity enriched MOA proteins (hierarchically ranked by the CoMOAenrich score) with the **MEDICASCY** top 100 MOA proteins (ranked by p-value), 88% of the diseases have proteins with a significant overlap p-value. The cumulative distribution for the CoMOAenrich scores and p-values for all the comorbidity enriched MOA proteins and the top 100 are shown in Figures S7 and S8, respectively. The summary statistics of the scores for these thresholds are shown in Table S1. For the comorbidity enriched MOA proteins ranked by their CoMOAenrich score, 58% have a p-value < 0.05. However, if one only assesses the top 100 comorbidity enriched MOA proteins, 94% have a p-value < 0.05, which are the proteins used for the global pathway analysis. Of the top 100 proteins used for pathway analysis, 82% have a p-value <0.005.

### Mapping of the LeMeDISCO MOA proteins to significant pathways

The cumulative distribution of the p-values for the pathways and the top 100 are shown in Figure S9 and the summary statistics are provided in Table S1. As shown in Figure S9, 62% of the pathways have a p-value < 0.015. We further note that there are some MOA proteins (e.g., SF3B1, BTAF1, and FAM160A1) and pathways (e.g., the nuclear receptor transcription pathway, SUMOylation of intracellular receptors, and PP2A-mediated dephosphorylation of key metabolic factors) that are enriched in approximately a third of the diseases in our library. This implies that there are homogenous molecular features across a subset of complex diseases. This has significant implications to disease interrelationships that will be explored elsewhere.

### Applications of LeMeDISCO

By way of illustration, we applied **LeMeDISCO** to two disparate diseases, coronary artery disease (CAD) and ovarian cancer (OC).

### Coronary artery disease (CAD)

CAD, a leading cause of death worldwide, is caused by narrowed or blocked arteries due to plaques composed of cholesterol or other fatty deposits lining the inner wall of the artery. These plaques result in decreased blood supply to the heart^17^. We find 2,747 significant comorbid diseases (p-value < 0.05), 1,459 comorbidity enriched MOA proteins (score > 0.01, meaning that at least one of the top 100 comorbid disease shares the protein as a MOA protein. This is the p-value weighted comorbidity frequency normalized by the number of comorbid diseases used for calculating the frequency. See Methods for more details. 7 significant pathways are associated with the top ranked 100 proteins (p-value < 0.05). The top 20 disease comorbidities, top 20 comorbidity enriched MOA proteins, and top 20 significant pathways are shown in Table 2. There are several cardiovascular-related significant comorbidities such as cardiovascular system disease, and myocardial infarction. Kidney disease, diabetes, obstructive lung disease and Alzheimer’s disease are also in the top ten with known comorbidities to CAD. Furthermore meta-analysis indicates an association between CAD and asthma, particularly in females with adult-onset asthma^19^. Prostanoid ligand receptors is the third most significant pathway found for CAD, which may be due to the number of COX-related comorbidity enriched proteins found. COX are involved in the synthesis of prostanoids. Prostanoids are structurally like lipids and are involved in thrombosis and other undesirable cardiovascular events^20^. CAD is also known to be comorbid with proteinuria, Alport syndrome, glomerulonephritis, liver disease and mitral valve insufficiency.

**Table 2.** Top 20 comorbidities (excluding same disease pair, (i.e. CAD-CAD)), top 20 comorbidity enriched MOA proteins (with respect to original disease), and top 20 (max) pathways associated with the prediction CAD results.

| Comorbidities |  |  | MOA proteins |  | Pathways |  |  |
| --- | --- | --- | --- | --- | --- | --- | --- |
| Disease | J -score | p-value | Gene name | Score | Pathway | Top pathway | p-value |
| Cardiovascular system disease | 0.49 | < 0.0001 | OSBPL8 | 0.38 | Nuclear Receptor transcription pathway | Gene expression (Transcription) | 4.60x10<sup>-14</sup> |
| Myocardial infarction | 0.46 | < 0.0001 | NR4A3 | 0.38 | SUMOylation of intracellular receptors | Metabolism of proteins | 0.01 |
| Heart disease | 0.45 | < 0.0001 | LXN | 0.38 | Prostanoid ligand receptors | Signal transduction | 0.01 |
| Kidney disease | 0.44 | < 0.0001 | SLC8A3 | 0.38 | Synthesis of very long-chain fatty acyl-CoAs | Metabolism | 0.03 |
| Diabetes mellitus | 0.44 | < 0.0001 | KCNA10 | 0.37 | Noncanonical activation of NOTCH3 | Signal transduction | 0.03 |
| Familial hyperlipidemia | 0.4 | < 0.0001 | NR3C2 | 0.37 | PP2A-mediated dephosphorylation of key metabolic factors | Metabolism | 0.04 |
| Congestive heart failure | 0.36 | < 0.0001 | OSBPL5 | 0.37 | E2F mediated regulation of DNA replication | Cell cycle | 0.05 |
| Obstructive lung disease | 0.35 | < 0.0001 | RARRES1 | 0.37 |  |  |  |
| Hepatorenal syndrome | 0.34 | < 0.0001 | COX7A2 | 0.36 |  |  |  |
| Alzheimer's disease | 0.34 | < 0.0001 | COX7A2L | 0.36 |  |  |  |
| Syndrome | 0.34 | < 0.0001 | COX7A1 | 0.36 |  |  |  |
| Glucose intolerance | 0.33 | < 0.0001 | GRP | 0.36 |  |  |  |
| Asthma | 0.33 | < 0.0001 | TSPAN13 | 0.36 |  |  |  |
| Proteinuria | 0.32 | < 0.0001 | ELOVL7 | 0.36 |  |  |  |
| Atherosclerosis | 0.32 | < 0.0001 | PLEKHG4 | 0.35 |  |  |  |
| Alport syndrome | 0.32 | < 0.0001 | HRASLS5 | 0.35 |  |  |  |
| Glomerulonephritis | 0.32 | < 0.0001 | PLA2G16 | 0.35 |  |  |  |
| Liver disease | 0.31 | < 0.0001 | ELOVL1 | 0.34 |  |  |  |
| Mitral valve insufficiency | 0.31 | < 0.0001 | RARRES3 | 0.34 |  |  |  |
| IgA glomerulonephritis | 0.31 | < 0.0001 | NR3C1 | 0.34 |  |  |  |

The above results were obtained without any extrinsic knowledge of CAD. Next, we show how additional information can be used. A GWAS study identified 155 CAD associated genes^21^. These GWAS genes associated with CAD were then used as input to GWAS-driven **LeMeDISCO**. The top 20 disease comorbidities, top 20 comorbidity enriched MOA proteins, and top 20 pathways are shown in Table 3. There were 136 predicted significant comorbidities (p-value < 0.05) by **LeMeDISCO**. There were 3,039 comorbidity enriched MOA proteins (score > 0.01) and 57 significant pathways (p-value < 0.05) found from global pathway analysis of the top 100 comorbidity enriched MOA proteins. The top comorbidities are anuria and renal artery disease, both associated with dysfunction of the kidneys. Anuria is attributed to failure of the kidneys to produce urine, and renal artery disease occurs when the arteries that supply blood and oxygen to the kidneys narrows. A study found an increase in renal artery stenosis in patients with CAD^22^. Other diseases that have at least some literature evidence of comorbidity with CAD are leukopenia, leukocyte disease, hereditary hemorrhagic telangiectasia, recurrent hypersomnia, lactic acidosis and possibly dilated cardiomyopathy. We were unable to find a link in the literature between CAD and skin hemangioma, plasma cell neoplasm, Xeroderma pigmentosum.

**Table 3.** Top 20 comorbidities, top 20 comorbidity enriched MOA proteins (with respect to original disease), and top 20 (max) pathways associated with the prediction CAD GWAS-driven **LeMeDISCO** results using the gene set from ^21^.

| Comorbidities |  |  | MOA proteins |  | Pathways |  |  |
| --- | --- | --- | --- | --- | --- | --- | --- |
| Disease | J-score | p-value | Gene name | Score | Pathway | Top pathway | p-value |
| Anuria | 0.04 | < 0.0001 | ZMYM6 | 0.09 | ERBB2 Activates PTK6 Signaling | Signal transduction | 7.10x10<sup>-6</sup> |
| Renal artery disease | 0.03 | 1.10E-10 | TARBP2 | 0.08 | ERBB2 Regulates Cell Motility | Signal transduction | 9.60x10<sup>-6</sup> |
| Skin hemangioma | 0.02 | 2.75E-05 | AFTPH | 0.08 | PI5P, PP2A and IER3 Regulate PI3K/AKT Signaling | Signal transduction | 0.0001 |
| Leukopenia | 0.02 | 0.0001 | MRPS12 | 0.08 | Signaling by ERBB2 TMD/JMD mutants | Disease | 0.0001 |
| Plasma cell neoplasm | 0.02 | 0.0002 | SYNM | 0.08 | Negative regulation of the PI3K/AKT network | Signal transduction | 0.0002 |
| Leukocyte disease | 0.02 | 0.0008 | EHBP1 | 0.07 | GRB2 events in ERBB2 signaling | Signal transduction | 0.0002 |
| Hereditary hemorrhagic telangiectasia | 0.02 | 0.002 | SGOL1 | 0.07 | PI3K events in ERBB2 signaling | Signal transduction | 0.0002 |
| Dilated cardiomyopathy | 0.02 | 0.003 | AFF2 | 0.07 | Signaling by ERBB2 KD Mutants | Disease | 0.0003 |
| Xeroderma pigmentosum | 0.01 | 0.003 | CLSTN3 | 0.07 | Signaling by ERBB2 in Cancer | Disease | 0.0003 |
| Recurrent hypersomnia | 0.01 | 0.003 | CD248 | 0.07 | Downregulation of ERBB2 signaling | Signal transduction | 0.0003 |
| Lactic acidosis | 0.01 | 0.005 | LRRCC1 | 0.07 | SHC1 events in ERBB2 signaling | Signal transduction | 0.0003 |
| Gastric antral vascular ectasia | 0.01 | 0.007 | PRICKLE3 | 0.07 | Constitutive Signaling by Aberrant PI3K in Cancer | Disease | 0.0008 |
| POEMS syndrome | 0.01 | 0.006 | MDFIC | 0.07 | TFAP2 (AP-2) family regulates transcription of growth factors and their receptors | Gene expression (Transcription) | 0.002 |
| Hemoglobin D disease | 0.01 | 0.006 | PCDH7 | 0.07 | Activation of TRKA receptors | Signal transduction | 0.002 |
| Hereditary spherocytosis | 0.01 | 0.006 | MOB3C | 0.07 | Downregulation of ERBB4 signaling | Signal transduction | 0.003 |
| Myelophthisic anemia | 0.01 | 0.006 | LOC101060321 | 0.07 | PI3K/AKT Signaling in Cancer | Disease | 0.004 |
| Protein-deficiency anemia | 0.01 | 0.006 | FAM8A1 | 0.07 | TRKA activation by NGF | Signal transduction | 0.007 |
| Hemolytic-uremic syndrome | 0.01 | 0.006 | LRRC17 | 0.06 | Signaling by ERBB2 | Signal transduction | 0.007 |
| Shwachman-Diamond syndrome | 0.01 | 0.006 | LRRTM2 | 0.06 | Signaling by Non-Receptor Tyrosine Kinases | Signal transduction | 0.009 |
| Favism | 0.01 | 0.006 | DISC1 | 0.06 | Signaling by PTK6 | Signal transduction | 0.009 |

ZMYM6, the top comorbidity enriched MOA protein, is highly enriched in the heart, skeletal muscle, kidney and liver, which is reflected in the type of disease comorbidities predicted. Further, there are two tropomyosin receptor kinase A (TRKA)-related pathways found via pathway analysis, activation of TRKA receptors and TRKA activation by nerve growth factor (NGF), a neurotrophin. Neurotrophins and their receptors (i.e. TRK) are known to have essential cardiovascular functions and may be suitable therapeutic targets^23^. With a score > 0.01, there are 1,459 and 3,039 comorbidity enriched MOA proteins from the **MEDICASCY** and GWAS-driven results, respectively. From this, 212 MOA proteins overlap, yielding a significant p-value (p-value < 0.0001). Thus, there is a significant overlap p-value from the GWAS driven **LeMeDISCO** results to the **MEDICASCY** MOA protein driven **LeMeDISCO** results for the predicted MOA proteins. **LeMeDISCO** does prioritize the most putatively important ones.

### Ovarian cancer (OC)

**LeMeDISCO** predicts 1,896 significant comorbidities to OC (p-value < 0.05), with 2,949 comorbidity enriched MOA proteins (score > 0.01). There were 67 significant pathways (p-value < 0.05) from the top 100 comorbidity enriched MOA proteins. The top 20 disease comorbidities, top 20 comorbidity enriched MOA proteins, and all significant pathways are shown in Table 4. The top comorbid disease associated with OC is thyroid gland cancer. Studies have found that OC can metastasize to the thyroid gland. There are several other high confidence cancers predicted to be comorbid with OC. Colorectal cancer and OC cooccur, and there have been a limited number of examples of cooccurrence of esophageal and pancreatic cancer with OC. OC from lung cancer metastasis occurs in <4% of OC patients. OC is found to be comorbid with stomach cancer and colon cancer, as are fallopian tube cancer and OC. Bile duct cancer is a very rare site of OC metastases. Metastatic OC spreads to the liver. Peritoneal cancer behaves similarly to OC. We were unable to find any literature support for the comorbidity of OC and urinary bladder cancer, although OC can affect urination. Nor is there literature support for the cooccurrence of OC with uterus interstitial leiomyoma, hemangioma of intra-abdominal structure, submucous uterine fibroid, cervical polyps, or lymphangioma.

**Table 4.**
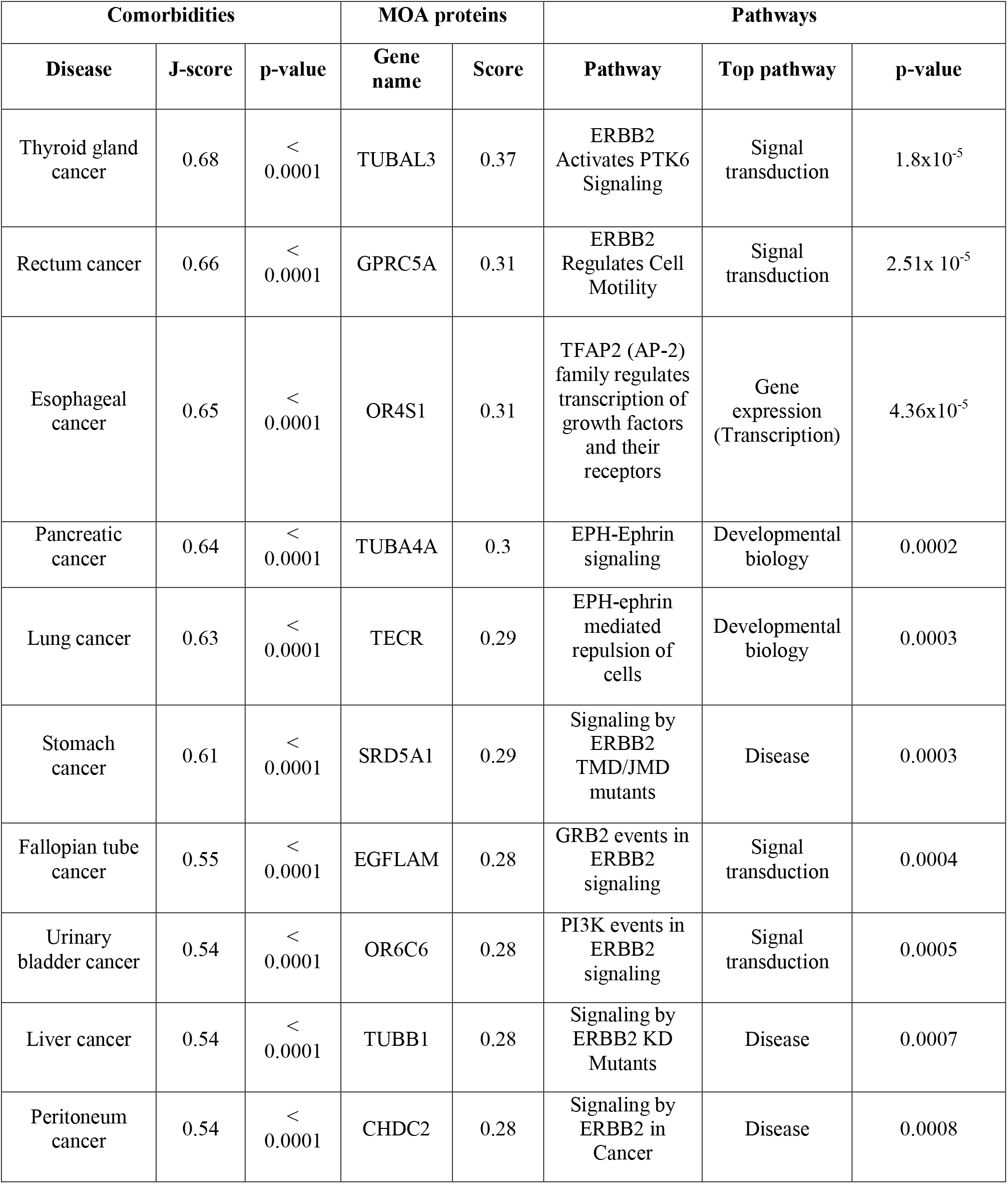
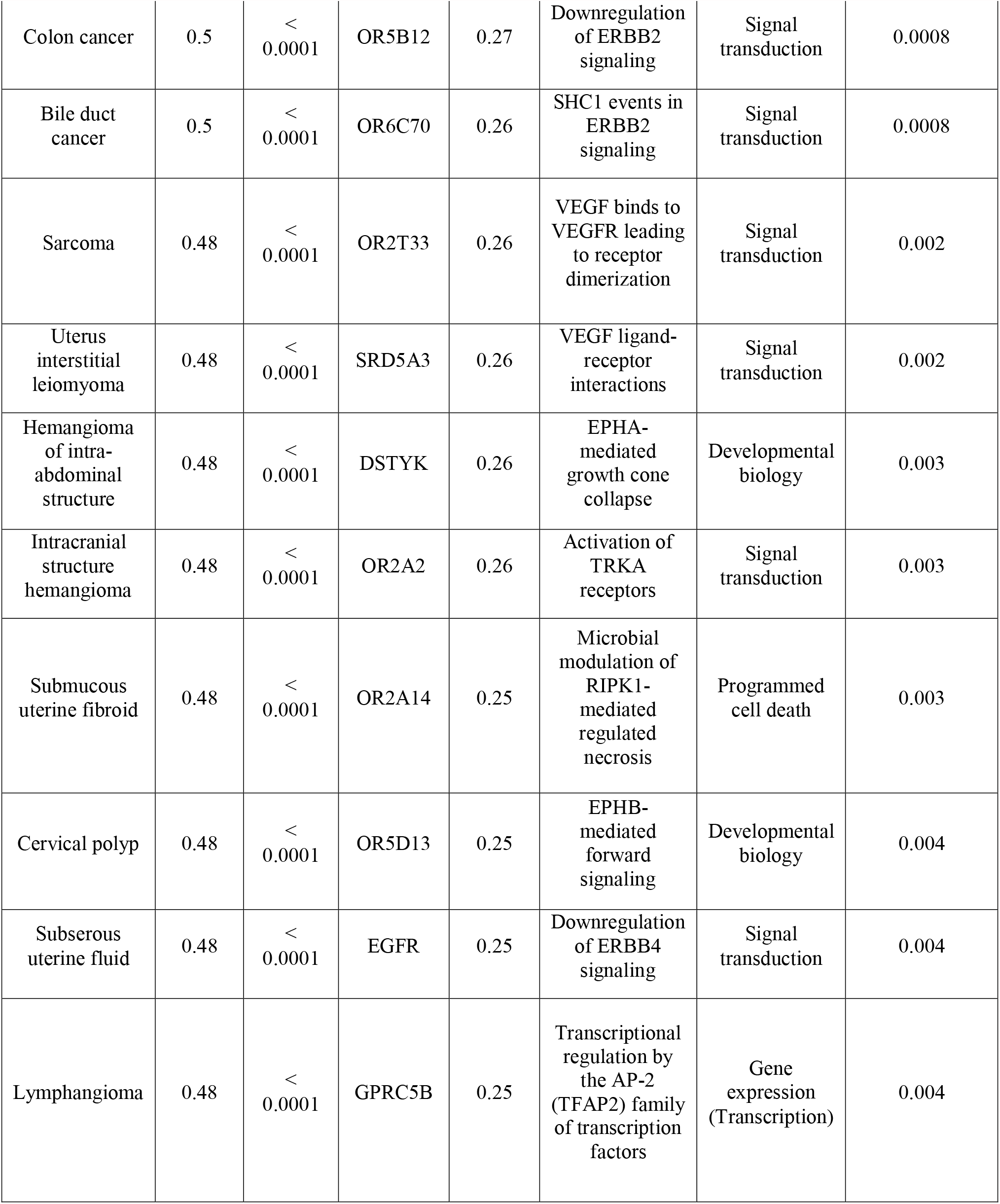
Top 20 comorbidities (excluding same disease pair, (i.e. OC-OC)), top 20 comorbidity enriched MOA proteins (with respect to original disease), and top 20 (max) pathways associated with the prediction OC results.

| Comorbidities |  |  | MOA proteins |  | Pathways |  |  |
| --- | --- | --- | --- | --- | --- | --- | --- |
| Disease | J-score | p-value | Gene name | Score | Pathway | Top pathway | p-value |
| Thyroid gland cancer | 0.68 | < 0.0001 | TUBAL3 | 0.37 | ERBB2 Activates PTK6 Signaling | Signal transduction | $1.8 \times 10^{-5}$ |
| Rectum cancer | 0.66 | < 0.0001 | GPRC5A | 0.31 | ERBB2 Regulates Cell Motility | Signal transduction | $2.51 \times 10^{-5}$ |
| Esophageal cancer | 0.65 | < 0.0001 | OR4S1 | 0.31 | TFAP2 (AP-2) family regulates transcription of growth factors and their receptors | Gene expression (Transcription) | $4.36 \times 10^{-5}$ |
| Pancreatic cancer | 0.64 | < 0.0001 | TUBA4A | 0.3 | EPH-Ephrin signaling | Developmental biology | 0.0002 |
| Lung cancer | 0.63 | < 0.0001 | TECR | 0.29 | EPH-ephrin mediated repulsion of cells | Developmental biology | 0.0003 |
| Stomach cancer | 0.61 | < 0.0001 | SRD5A1 | 0.29 | Signaling by ERBB2 TMD/JMD mutants | Disease | 0.0003 |
| Fallopian tube cancer | 0.55 | < 0.0001 | EGFLAM | 0.28 | GRB2 events in ERBB2 signaling | Signal transduction | 0.0004 |
| Urinary bladder cancer | 0.54 | < 0.0001 | OR6C6 | 0.28 | PI3K events in ERBB2 signaling | Signal transduction | 0.0005 |
| Liver cancer | 0.54 | < 0.0001 | TUBB1 | 0.28 | Signaling by ERBB2 KD Mutants | Disease | 0.0007 |
| Peritoneum cancer | 0.54 | < 0.0001 | CHDC2 | 0.28 | Signaling by ERBB2 in Cancer | Disease | 0.0008 |
| Colon cancer | 0.5 | < 0.0001 | OR5B12 | 0.27 | Downregulation of ERBB2 signaling | Signal transduction | 0.0008 |
| Bile duct cancer | 0.5 | < 0.0001 | OR6C70 | 0.26 | SHC1 events in ERBB2 signaling | Signal transduction | 0.0008 |
| Sarcoma | 0.48 | < 0.0001 | OR2T33 | 0.26 | VEGF binds to VEGFR leading to receptor dimerization | Signal transduction | 0.002 |
| Uterus interstitial leiomyoma | 0.48 | < 0.0001 | SRD5A3 | 0.26 | VEGF ligand-receptor interactions | Signal transduction | 0.002 |
| Hemangioma of intra-abdominal structure | 0.48 | < 0.0001 | DSTYK | 0.26 | EPHA-mediated growth cone collapse | Developmental biology | 0.003 |
| Intracranial structure hemangioma | 0.48 | < 0.0001 | OR2A2 | 0.26 | Activation of TRKA receptors | Signal transduction | 0.003 |
| Submucous uterine fibroid | 0.48 | < 0.0001 | OR2A14 | 0.25 | Microbial modulation of RIPK1-mediated regulated necrosis | Programmed cell death | 0.003 |
| Cervical polyp | 0.48 | < 0.0001 | OR5D13 | 0.25 | EPHB-mediated forward signaling | Developmental biology | 0.004 |
| Subserous uterine fluid | 0.48 | < 0.0001 | EGFR | 0.25 | Downregulation of ERBB4 signaling | Signal transduction | 0.004 |
| Lymphangioma | 0.48 | < 0.0001 | GPRC5B | 0.25 | Transcriptional regulation by the AP-2 (TFAP2) family of transcription factors | Gene expression (Transcription) | 0.004 |

Tubulin Alpha Like 3 (TUBAL3) is the topmost comorbidity enriched MOA protein. Tubulin proteins are associated with breast cancer, which can co-occur with OC^24^. There are also enriched pathways associated with ephrin ligands. Aggressive forms of ovarian cancer have been previously investigated to upregulate forms of ephrin, such as ephrinA5^25^. There are 14 ephrin-related comorbidity enriched MOA proteins found (score > 0.05)^26^.

We next examined a set of 11 genes associated with OC risk from a study that assessed the multiple-gene germline sequences in 95,561 women with OC into **LeMeDISCO**^27^. The results for the top 20 comorbidities, MOA proteins, and pathways associated are shown in in Table 5. There were 207 significant comorbidities (p-value < 0.05) predicted, 2,895 comorbidity enriched MOA proteins and 5 significant pathways associated with the top ranked 100 proteins (p-value < 0.05). The top hit comorbidity associated with OC was Sertoli-Leydig cell tumor, a rare cancer of the ovaries, which can yield an increase in the male sex hormone, testosterone^28^. Sex cord-gonadal stromal tumor is a rare type of ovarian cancer. There is some evidence for the comorbidity of diffuse scleroderma, severe acute respiratory syndrome, hyperuricemia, coronary stenosis, lymphatic system disease, germinoma, embryonal cell carcinoma and OC. Germinoma, another comorbidity predicted to be associated with OC, is a tumor often found in the brain is typically formed due to dysfunctional localization of germ cells to their respective locations. Furthermore, hemoglobinopathy, a disease(s) of the blood, was also a comorbidity associated with OC. A study found a relationship between hemoglobin levels and interleukin-6 levels in individuals with untreated epithelial ovarian cancer, which indicated the inflammatory role in cancer-associated anemia^29^. There is scant literature evidence for the comorbidity of adenosquamous carcinoma or acinar cell carcinoma with OC.

**Table 5.** Top 20 comorbidities, top 20 comorbidity enriched MOA proteins (with respect to original disease), and top 20 (max) pathways associated with the prediction OC GWAS driven results using the gene set from ^27^.

| Comorbidities |  |  | MOA proteins |  | Pathways |  |  |
| --- | --- | --- | --- | --- | --- | --- | --- |
| Disease | J-score | p-value | Gene name | Score | Pathway | Top pathway | p-value |
| Sertoli-Leydig cell tumor | 0.004 | 7.11E-05 | GMPR | 0.23 | RAB geranylgeranylation | Metabolism of proteins | 7.88x10<sup>-8</sup> |
| Sex cord-gonadal stromal tumor | 0.004 | 7.11E-05 | TUBE1 | 0.23 | RAB regulation of trafficking | Vesicle-mediated transport | 0.003 |
| Diffuse scleroderma | 0.004 | 2.59E-05 | NTPCR | 0.23 | RAB GEFs exchange GTP for GDP on RABs | Vesicle-mediated transport | 0.005 |
| Severe acute respiratory syndrome | 0.004 | 2.65E-05 | AK4 | 0.23 | Interconversion of nucleotide di- and triphosphates | Metabolism | 0.008 |
| Hyperuricemia | 0.004 | 3.82E-05 | AK2 | 0.23 | TBC/RABGAPs | Vesicle-mediated transport | 0.01 |
| Coronary stenosis | 0.004 | 1.31E-05 | ASMTL | 0.23 |  |  |  |
| Lymphatic system disease | 0.004 | 0.0006 | RAB35 | 0.23 |  |  |  |
| Adenosquamous carcinoma | 0.003 | 7.95E-05 | AK6 | 0.22 |  |  |  |
| Germinoma | 0.003 | 0.002 | TUBD1 | 0.22 |  |  |  |
| Hemoglobinopathy | 0.003 | 2.73E-05 | OAS2 | 0.22 |  |  |  |
| Genetic disease | 0.003 | 3.11E-05 | RRM1 | 0.22 |  |  |  |
| Embryonal cell carcinoma | 0.003 | 0.0003 | TUT1 | 0.22 |  |  |  |
| Acinar cell carcinoma | 0.003 | 0.0003 | IDNK | 0.22 |  |  |  |
| Inflammatory breast carcinoma | 0.003 | 0.0001 | ARL13B | 0.22 |  |  |  |
| Biliary tract disease | 0.003 | 0.0004 | SMG9 | 0.22 |  |  |  |
| Fanconi anemia | 0.003 | 4.14E-05 | ARL5A | 0.22 |  |  |  |
| Ovarian carcinoma | 0.003 | 0.0004 | ERG | 0.22 |  |  |  |
| Uveal cancer | 0.003 | 1.39E-05 | GTPBP8 | 0.22 |  |  |  |
| Carcinosarcoma | 0.003 | 0.0002 | GIMAP8 | 0.22 |  |  |  |
| Salivary gland adenoid cystic carcinoma | 0.003 | 0.0005 | GNL3 | 0.22 |  |  |  |

One of the top comorbidity enriched MOA proteins found is GMPR, also found to be up-regulated in metastatic serous papillary ovarian tumors from a differential gene expression analysis^30^. RAB-related pathways are provided by the pathway analysis. Rab35, a protein associated with modification of actin remodeling^31^, is a top 20 comorbidity enriched MOA protein that has been shown to be upregulated in individuals with OC under androgen treatment. Notably, there was a significant overlap p-value from the GWAS driven **LeMeDISCO** results to the **MEDICASCY** MOA protein driven **LeMeDISCO** results for the predicted comorbidities (p-value = 5.3×10^−12^) and MOA proteins (p-value < 0.0001).

### LeMeDISCO web server

The **LeMeDISCO** web service allows researchers to query our library of 3,608 diseases or input a set of pathogenic human genes/proteins and compute their predicted comorbidities, MOA proteins, and pathways associated. The web service is freely available for academic users at http://sites.gatech.edu/cssb/LeMeDISCO.

## Discussion

**LeMeDISCO** is a systematic approach for studying and analyzing possible features underlying the common proteins underlying a set of comorbid diseases. The resulting predicted driver proteins and pathways for each disease or input gene set can allow researchers to generate new diagnostic and treatment options and hypotheses. Interestingly, there were some MOA proteins and pathways present across approximately a third of the diseases, implying common disease drivers. The implications of this observation and its relationship to disease origins will be pursued in future work. We do note that the current comorbid disease analysis strongly suggests that the *“one target-one disease-one molecule”* approach often used in developing disease therapeutics^32^ is likely somewhat too simplistic.

To fully understand the complexities of a disease, one must trace the origin of its pathogenesis, which may be due to a variant that is somehow related to the condition. However, such variants may also be associated with a disease not previously known to be associated with that condition. Such interrelations can be further investigated by identifying high confidence comorbidity predictions from **LeMeDISCO**, regardless of whether or not their comorbidity was previously known in the literature. For example, analysis of the comorbid diseases associated with CAD and OC have not only recapitulated known disease comorbidities but have also provided novel insights. The **MEDICASCY** and GWAS-driven results for CAD yielded high confidence associations between hepatic diseases and forms of anemia, which can be further investigated through the comorbidity enriched MOA proteins and pathways. Furthermore, the **MEDICASCY** and GWAS-driven results for OC revealed more high confidence associations to other forms of cancer such as thyroid gland cancer and Sertoli-Leydic cell tumor.

**LeMeDISCO** not only has applications to the study of the underlying etiology behind a disease but may also be used during the early stages of drug discovery to identify efficacious drugs. Rather than starting with a small molecule or protein target of choice, **LeMeDISCO** allows one to begin at the level of disease biology, often termed phenotypic drug discovery. In future work, we shall demonstrate the utility of **LeMeDISCO** in identifying efficacious drugs to treat a given disease. Overall, the results of the current analysis and preliminary applications to drug discovery suggest that **LeMeDISCO** provides a set of tools for elucidating disease etiology and interrelationships and that a more systems wide, comprehensive approach to both personalized medicine and drug discovery is required.

## Data Availability

The web service is freely available for academic users at http://sites.gatech.edu/cssb/LeMeDISCO.

## Acknowledgments

This project was funded by R35GM118039 of the Division of General Medical Sciences of the NIH. We thank Bartosz Ilkowski for internal computing support and Jessica Forness for proof-reading the manuscript.

## Author Contributions

CA, HY and JS conceived of the method; CA and HY implemented the method, CA, HY and JS analyzed the data and wrote the paper.

## Data availability

The web service is freely available for academic users at http://sites.gatech.edu/cssb/LeMeDISCO.

## Methods

### Overview of LeMeDISCO

A flowchart of **LeMeDISCO** is shown in Figure 2. **LeMeDISCO** employs **MEDICASCY**^11^ to predict possible disease MOA proteins. Here, **MEDICASCY** is applied in prediction mode (i.e., any training drugs having a Tanimoto-Coefficient =1 to a given input drug is excluded from training) to avoid a strong bias towards drugs in the training set on a set of 2,095 FDA-approved drugs^32^. For each of the 3,608 indications, we rank the 2,095 probe drugs according to their Z-scores, Z_d_, defined using the raw score computed by **MEDICASCY** from:

**Figure 2.**
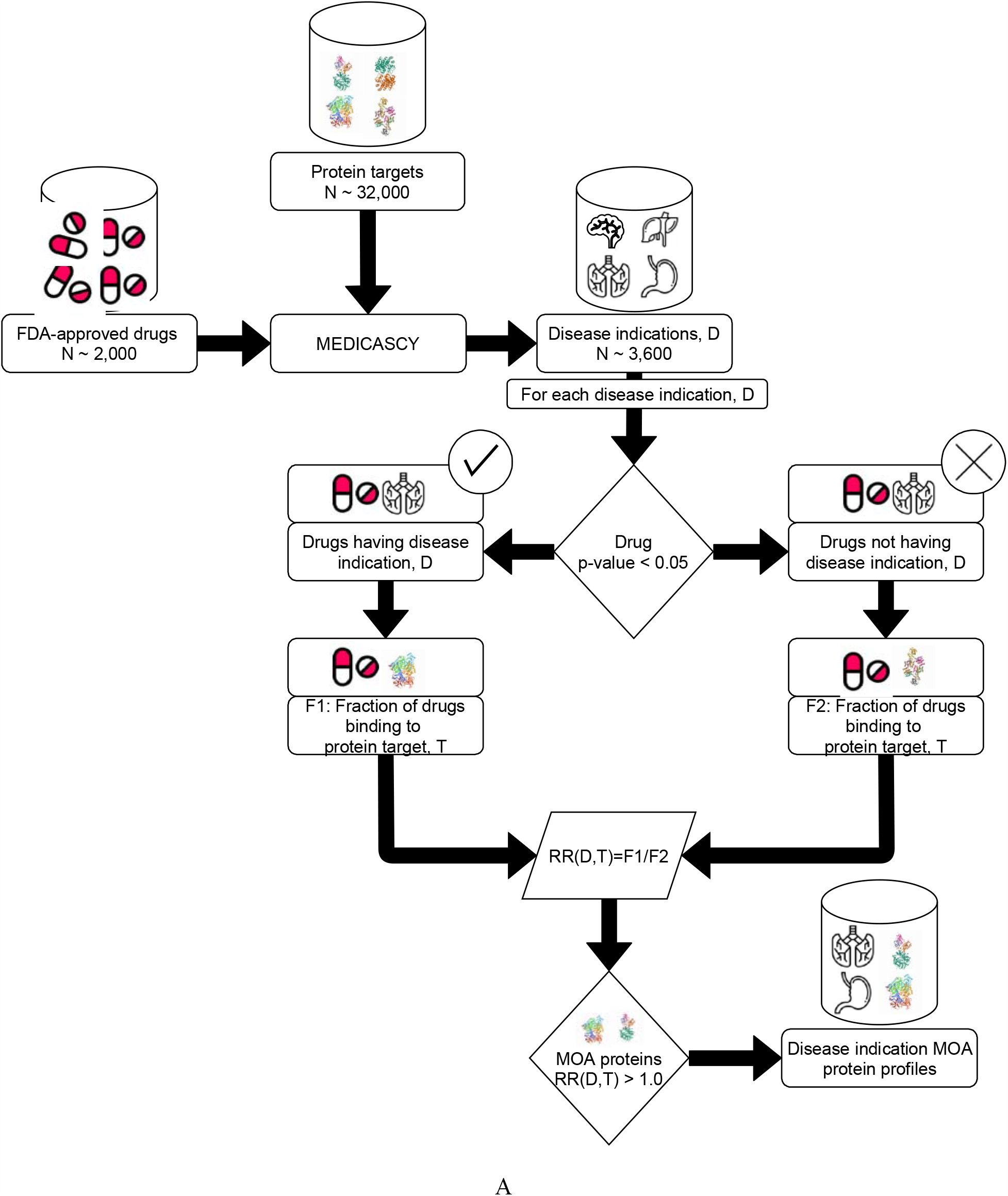
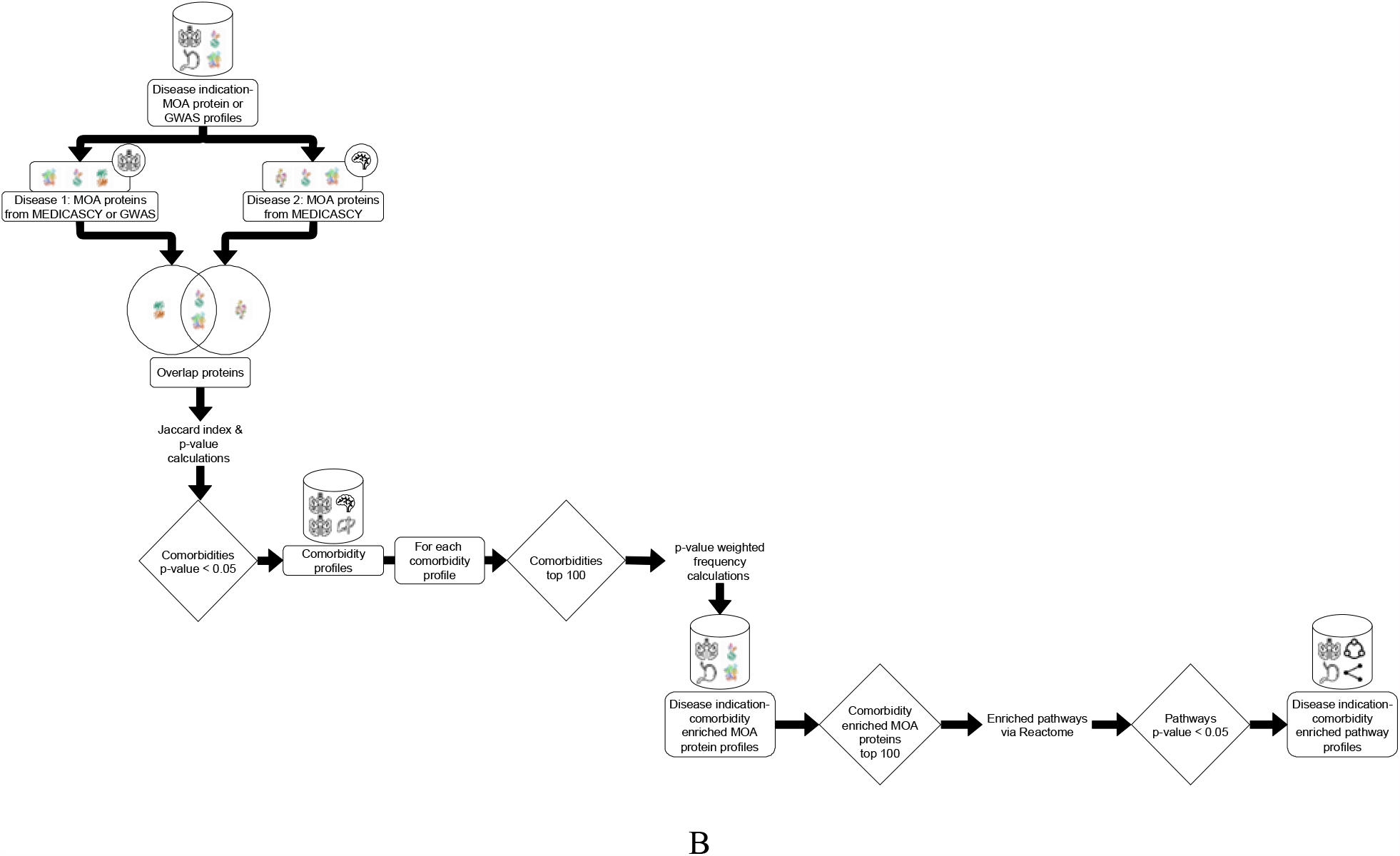
**LeMeDISCO** schematic representation of (A) the method for determining the MOA proteins associated with a disease indication via **MEDICASCY**, and (B) the method for determining the comorbidities associated with a given disease and its molecular mechanisms via LeMeDISCO.

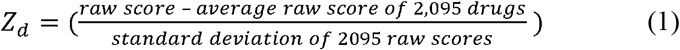

To predict a drug as having the given indication, we applied a Z_d_ cutoff of 1.65 that approximately corresponds to a p-value of 0.05 for the upper tailed null hypotheses of random Z_d_. Thus, for each indication D, the 2,095 probe drugs are separated into two groups: N_1_ are predicted to have indication D (Z_d_≥1.65) and N_2_ (=2,095-N_1_) are not predicted to have indication D (Z_d_<1.65). This is a very loose prediction of a drug’s indication with the advantage that it always predicts some drugs having the indication with its expected statistical confidence. Then, for a given indication D and each protein target, T, in the human proteome of our modeled 32,584 proteins, there are a subset of the drugs (or perhaps none) predicted by **FINDSITE**^**comb2.0** 33^ to bind T. The relative risk RR(D,T) of the given target T with respect to indication D as:

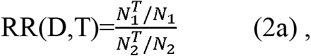

where 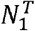 and 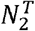 are the numbers of drugs binding to T with and without indication D, respectively. The numerator is the estimation of the probability of drugs having the predicted indication D (Z_d_≥1.65) that bind to protein 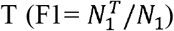. The denominator is the probability of finding drugs that do not have the predicted indication D but which bind to protein 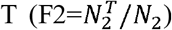. This latter probability serves as the background probability that an arbitrary drug will bind to T. When no drug is predicted to bind to protein T, RR(D,T) is set to zero. RR(D,T)=F1/F2 > 1 means that a drug having indication D is more likely to bind to T than arbitrary drugs not having the predicted indication D will bind to T.

We then compute the statistical significance of RR(D,T) by calculating a p-value using Fisher’s exact test^34,35^ on the following contingency table:

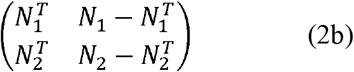

We define a protein target T as predicted to be a possible MOA target for indication D if its p-value < 0.05 because it is more likely to be targeted by efficacious drugs than arbitrary drugs. Thus, for each of the 3,608 indications, there is a list of predicted possible MOA proteins.

To reduce false positive MOAs, we utilized the human protein atlas database (https://www.proteinatlas.org/about/download, *normal_tissue*.*tsv*) of expression profiles for proteins in normal human tissues based on immunohistochemisty using tissue micro arrays^36^ to filter those proteins that are “not detected” and not “uncertain” in all tested tissues related to an indication. To determine the tissues related to an indication, tissues are mapped to their ICD-10 main codes and indications having the same main codes are related to the tissue.

Using the input of two sets of putative MOA proteins having a p-value of < 0.05 calculated by Fisher’s exact test^34^, we calculate their Jaccard index^37^ J(D_1_,D_2_) (J-score) defined in eq. 3a as

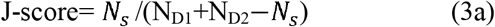

We then calculate the p-value for significance by Fisher’s exact test for the contingency table^34^ that gives the probability of having overlap ≥ *N*_*s*_ by randomly selecting *N*_*D*2_ out of *N*_*t*_ proteins^34,38^:

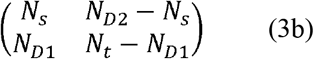

*N*_*D*1_, *N*_*D*2_ are the numbers of MOA proteins/genes of disease D_1_ and D_2_; *N*_*s*_ is the number of overlapped MOA proteins between D_1_, D_2_ and *N*_*t*_ is the total number of human proteins. The Jaccard index J-score is a statistical measure of the similarity between MOA proteins of D_1_ and D_2,_ and its value ranges between 0 and 1. Since the null hypothesis of *N*_*s*_ corresponds to a hypergeometric distribution, the p-value of observing the number of overlapped MOA proteins between D_1_, D_2_ ≥ *N*_*s*_ can be calculated using Fisher’s exact test on the table in eq. 3b^35^. We will use the J-score for predicting comorbidity and compare it with the observed comorbidity.

In large scale disease-disease comorbidity calculations, we use the MOAs predicted by **MEDICASCY**^11^. In addition, MOA targets between disease pairs can also be derived from experimental data; examples include differential gene expression (GE), Mendelian or somatic mutation profiles comparing disease vs. control normal samples, better vs. worse prognosis samples, or drug treated vs. control untreated samples^39^.

### Benchmarking of LeMeDISCO

We validated **LeMeDISCO**’s J-score by correlating it with the observed comorbidity as quantified by (a) the logarithm of relative risk log(RR) score and (b) the φ**-**score (Pearson’s correlation for binary variables)^14^. The relative risk (RR) is the probability that two diseases cooccur in a single individual relative to random. Since RR scales exponentially with respect to the strength of two interacting diseases, we use log(RR) for correlation analysis. The log(RR) and φ**-**score are computed from US Medicare insurance claim data using^14^:

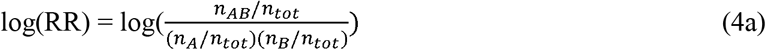

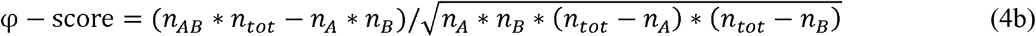

where n_tot_ = total number of patients; n_A_, n_B_ = number of patients diagnosed with disease A and B, and n_AB_ = number of patients diagnosed with both diseases A and B.

### Identification of key MOA proteins and associated pathways for disease comorbidity

After determining the significant comorbidities for each disease, the p-value weighted frequency of shared MOA proteins across the top 100 predicted comorbidities are calculated. We define a p-value weighted frequency of an input MOA as follows (i.e. CoMOAenrich score): If MOA protein T is shared by a comorbid indication D and the p-value of T associated with D is *P*, then the weight defined by the min(1.0,-αlog*P*) is counted as T’s frequency. In practice, we used 10 cancer cell line data^40^ to optimize the coefficient α to 0.025. We further computed a p-value via 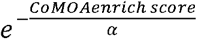 where α = 0.025, as previously mentioned. These MOA proteins expand the number of possible molecular players driving disease pathogenesis. An empirically derived CoMOAenrich score (normalized by the number of comorbid indications that is 100) threshold of 0.01 was used, which is equivalent to 1% of the comorbid indications having the MOA proteins with a significant p-value (<4.2 × 10^−18^). Then, up to the top 100 comorbidity enriched MOA proteins for each disease were used in global pathway analysis via Reactome^12^. The pathways with a p-value < 0.05 were extracted. The frequency of pathways across diseases was assessed to identify common pathways of disease.

### LeMeDISCO usage

As shown in Figure 2, **LeMeDISCO** can be used in two different ways: 1.) **MEDICASCY**-driven **LeMeDISCO**: The comorbidities for any of the 3,608 diseases from the **MEDICASCY** provided MOA proteins are predicted (Figure 2A). 2.) Pathogenic gene set driven **LeMeDISCO**: Input your own pathogenic gene set derived from differential gene expression, GWAS, exome analysis, or other experimental/clinical techniques (shown in Figure 2B). The **LeMeDISCO** web service allows users to query the **LeMeDISCO** database as well as input their own set of pathogenic genes to assess the associated comorbidities, MOA proteins, and pathways.

