## Supplemental Materials for "LeMeDISCO: A computational method for large-scale prediction & molecular interpretation of disease comorbidity"

**Supplementary Figures**

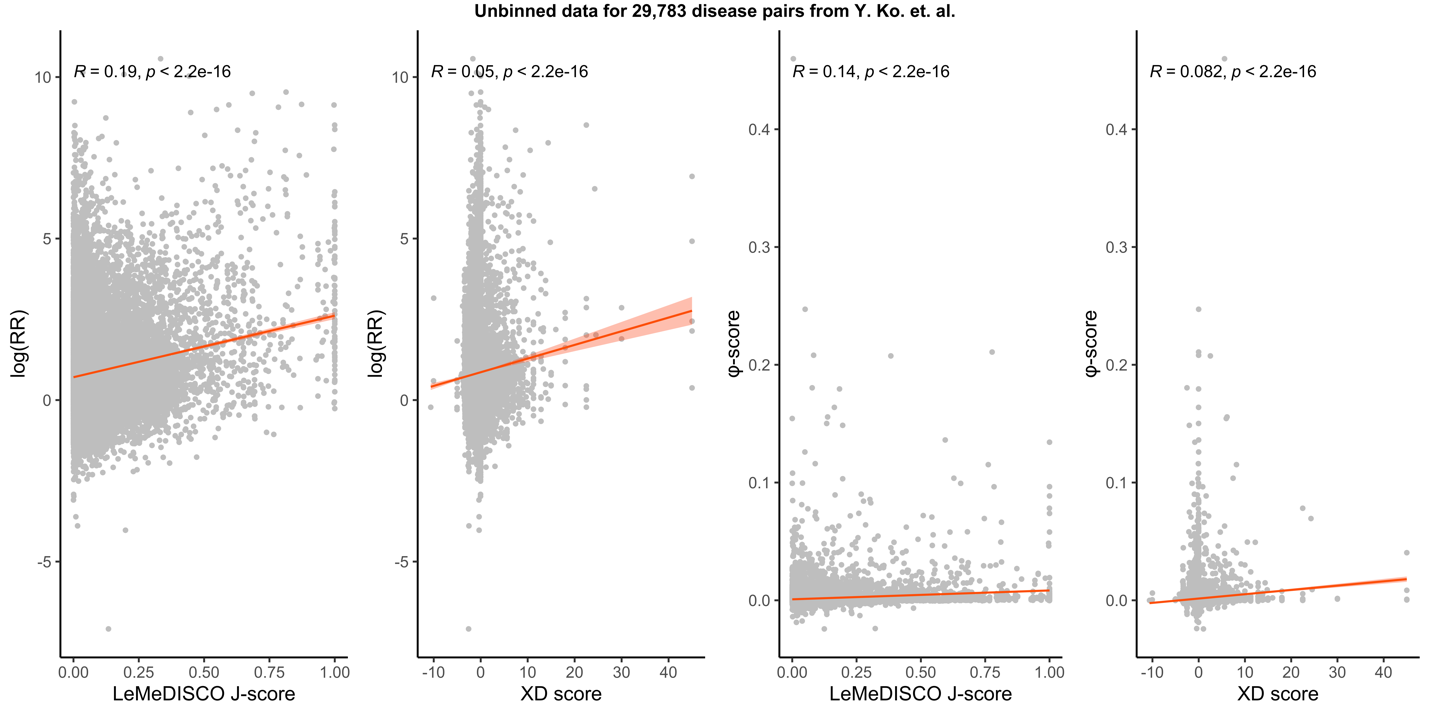

**Figure S1.** Correlation between the XD score^7^ and J-score to the log(RR) score and φ**-**score.

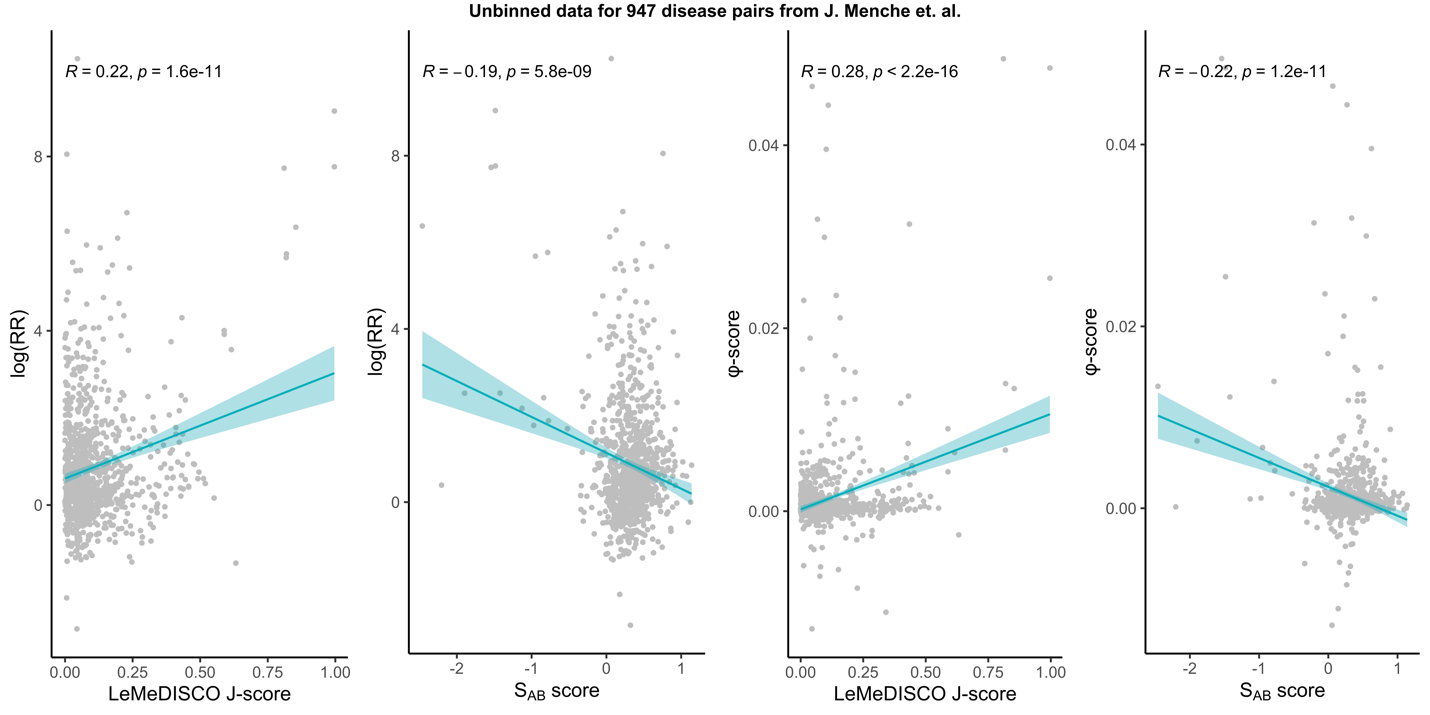
 **Figure S2.** The correlation between the *S_AB_* score ^6^ and J-score to the log(RR) score and φ**-**score.

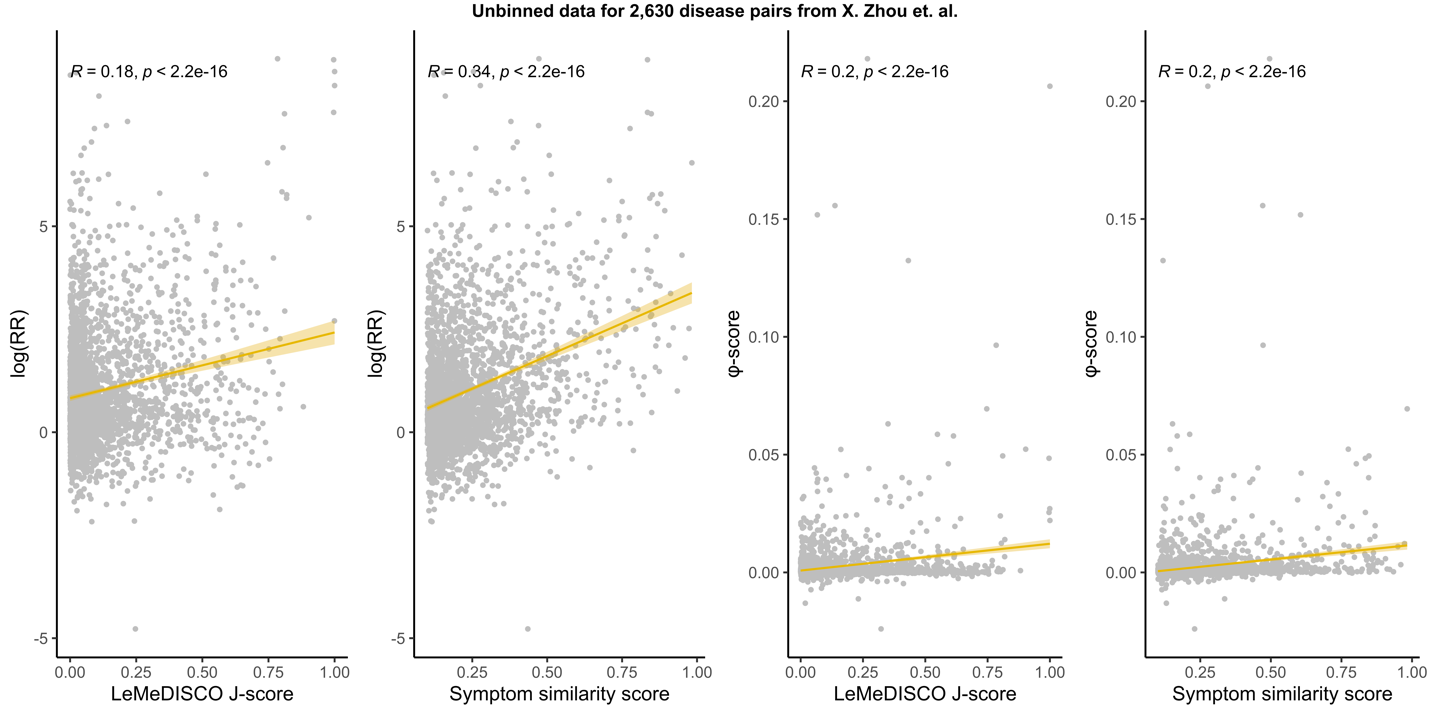

**Figure S3.** Correlation between the symptom similarity ^5^ and J-score to the log(RR) score and φ**-**score.

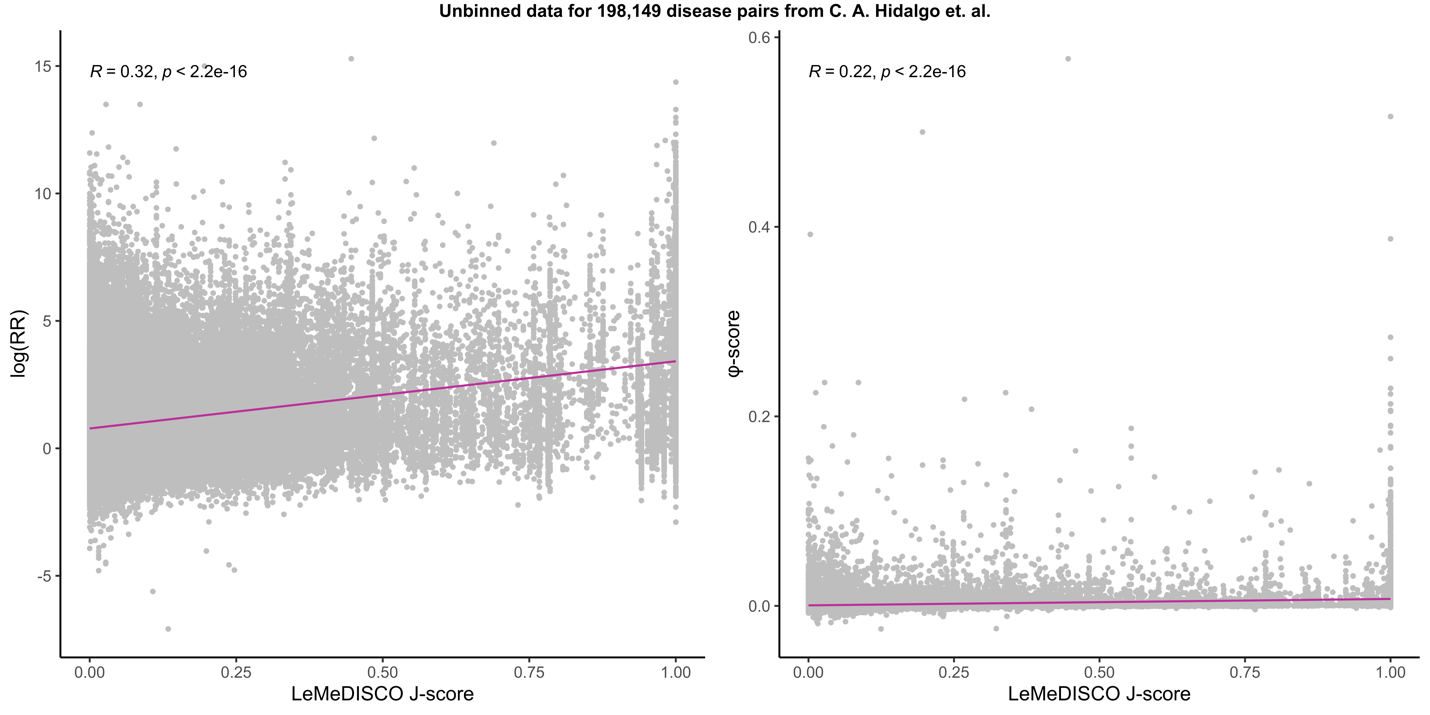

**Figure S4.** Correlation between the J-score to the log(RR) score and φ**-**score for the 198,149 diseases from ref^27^.

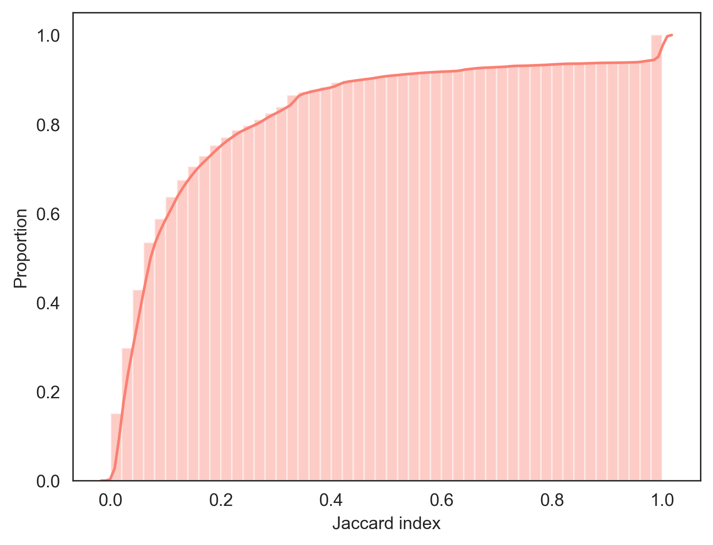

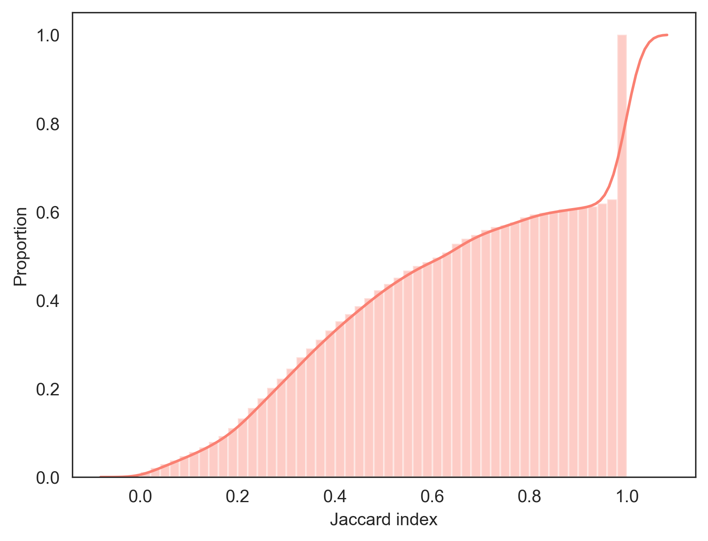

A	B

**Figure S5**. (A.) Cumulative distribution function (CDF) of the comorbidities with p-value < 0.05 and (B) the top 100 comorbidities hierarchically ranked by J-score with p-value < 0.05 (B.)

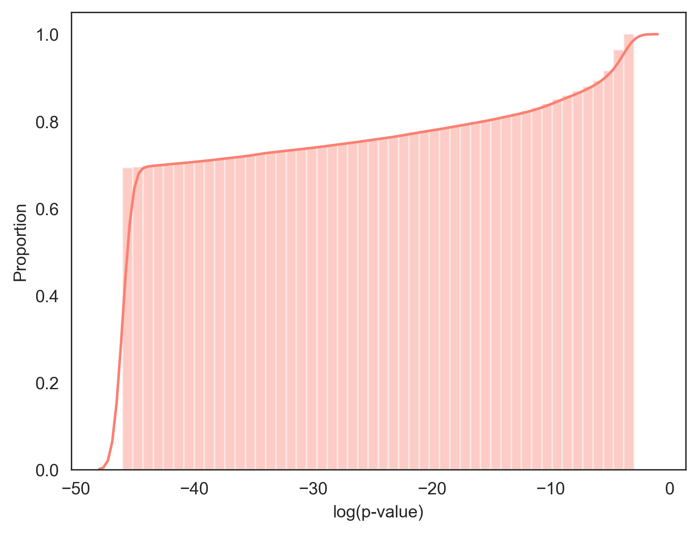

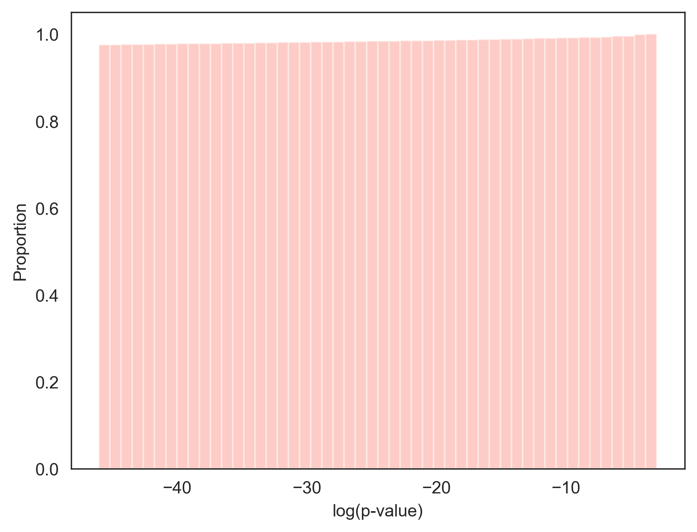

A	B

**Figure S6**. CDFs of the comorbidities’ log(p-values) with p-value < 0.05 (A.) and top 100 most comorbidities hierarchically ranked by J-score with p-value < 0.05 (B.). p-values <1.0x10^-20^ were set to1.0x10^-20^.

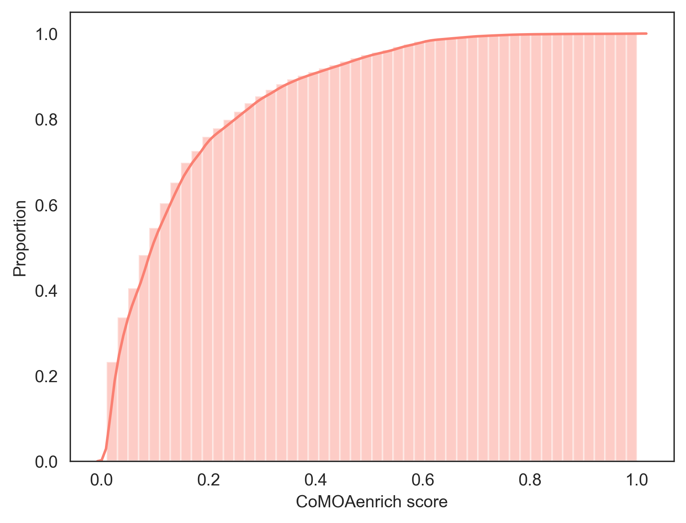

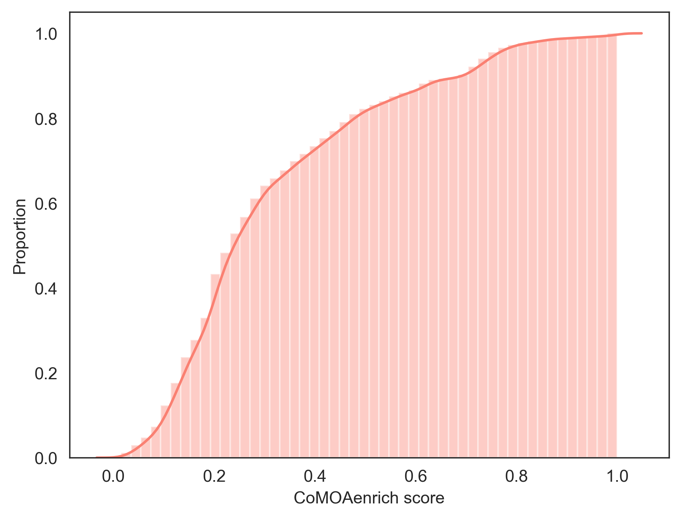

A	B

**Figure S7**. CDFs of (A). comorbidity enriched MOA protein CoMOAenrich scores with CoMOAenrich score > 0.01 and (B) the top the 100 comorbidity enriched MOA proteins hierarchically ranked by the CoMOAenrich score with CoMOAenrich score > 0.01.

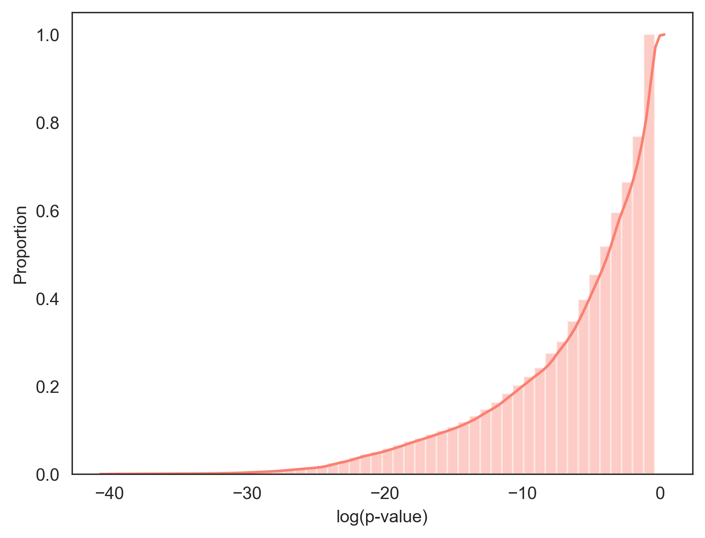

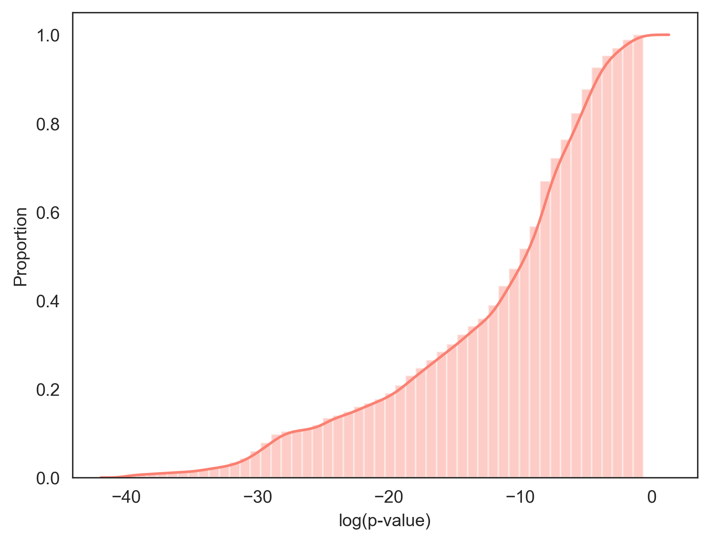

A	B

**Figure S8**. CDFs of (A) comorbidity enriched MOA protein log(p-values) with CoMOAenrich score > 0.01 (A.) and (B) top 100 comorbidity enriched MOA protein hierarchically ranked by the CoMOAenrich score with CoMOAenrich score > 0.01. p-values < 1.0x10^-20^ were set to1.0x10^-20^.

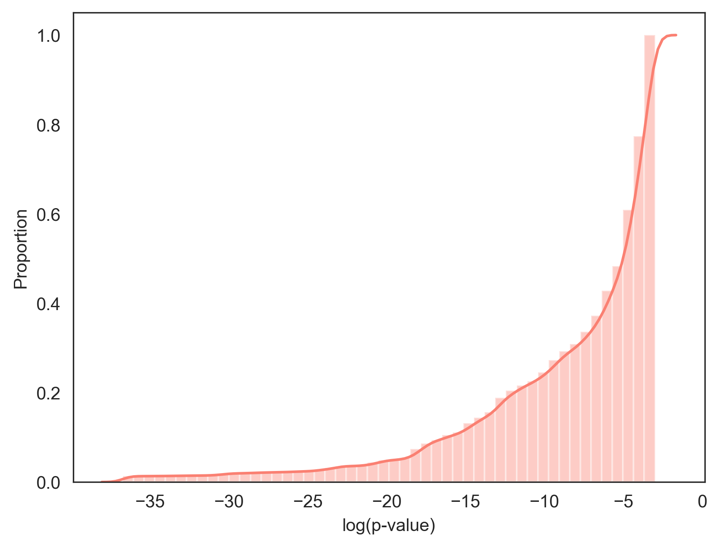

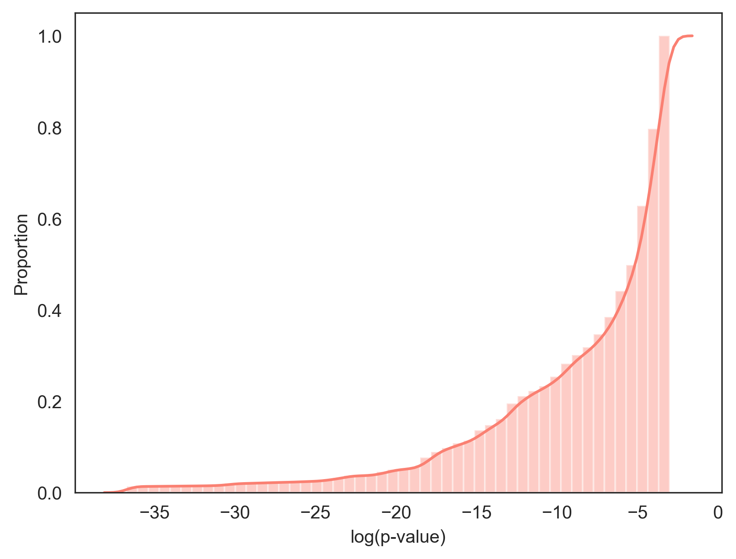

A	B

**Figure S9**. CDFs of (A). comorbidity enriched pathways log(p-values) with p-value < 0.05 and (B)top 100 comorbidity enriched pathways with p-value < 0.05 ranked by p-values in ascending order (B).

**Supplementary Tables**

**Table. S1.** Summary statistics of the MEDICASCY-driven LeMeDISCO scores for the comorbidities (excluding diagonals), comorbidity enriched MOA proteins, and pathways.

|  | **Count^a^** | **Mean^b^** | **Standard deviation^c^** | **Min^d^** | **Max^d^** |
| --- | --- | --- | --- | --- | --- |
| **Comorbidities** | | | | | |
| J-score | | | | | |
| p-value < 0.05 | 5,987,682 | 1.8e-01 | 2.5e-01 | 5.8e-04 | 1.0 |
| Top 100 &  p-value < 0.05 | 363,886 | 0.63 | 0.34 | 0.001 | 1.0 |
| p-value | | | | | |
| p-value < 0.05 | 5,987,682 | 2.2e-03 | 7.4e-03 | 0.0 | 4.99e-02 |
| Top 100 &  p-value < 0.05 | 363,886 | 8.0-05 | 1.26e-03 | 0.0 | 4.98e-02 |
| **Comorbidity-enriched MOA proteins** | | | | | |
| CoMOAenrich score | | | | | |
| CoMOAenrich score > 0.01 | 5,746,520 | 1.5e-01 | 1.57e-01 | 1.0e-02 | 1.0 |
| Top 100 &  CoMOAenrich score > 0.01 | 364,306 | 0.31 | 0.21 | 0.02 | 1.0 |
| p-value | | | | | |
| CoMOAenrich score > 0.01 | 5,746,520 | 1.5e-01 | 2.1e-01 | 4.2e-18 | 6.7e-01 |
| Top 100 & CoMOAenrich score > 0.01 | 364,306 | 1.07e-02 | 4.57e-02 | 4.2e-18 | 5.4e-01 |
| **Pathways** | | | | | |
| p-value | | | | | |
| p-value < 0.05 | 120,752 | 1.29e-02 | 1.45e-02 | 1.1e-16 | 4.99e-02 |
| Top 100 &  p-value < 0.05 | 117,090 | 1.21e-02 | 1.39e-02 | 1.1e-16 | 4.99e-02 |

^a^The count is the raw frequency of the comorbidities, MOA proteins, and pathways across diseases.

^b^The mean is the average of the respective scores/values across diseases.

^c^The standard deviation represents the variation of the scores/values across diseases.

^c^The min and max values are the absolute minimum and maximum score/value observed across diseases.
